# Diet Soda Consumption is Associated with Shifts in Gut Microbiome Function of Overweight Young Women

**DOI:** 10.64898/2026.11.12.26367968

**Authors:** Philip Y. Appiah, Marcos Peréz-Losada, Allison C. Sylvetsky, Keith A. Crandall, Carmen P. Ortega-Santos

## Abstract

**Background:** Low-calorie sweeteners (LCS) are consumed as sugar substitutes and may alter gut microbial composition. Their effects on microbial functional pathways and cardiometabolic physiology in humans remain poorly understood, particularly among individuals with overweight.

**Methods:** We conducted a secondary analysis of an 8-week, single-arm pre/post intervention involving eight overweight young adult women who consumed sucralose- and acesulfame-potassium-sweetened Diet Pepsi three times daily. Shotgun metagenomic sequencing of stool samples was used to profile microbial functional pathways with HUMAnN2. Multivariable linear mixed models (MaAsLin2) assessed associations between pathways and metabolic biomarkers. We hierarchically clustered nominally significant pathways into six representative groups and calculated Spearman correlations with interleukin-6 (IL-6), leptin, and HOMA-IR. P values were adjusted using the Benjamini-Hochberg false discovery rate (FDR) procedure. Biomarkers were measured using clinical chemistry and immunoassays.

**Results:** MaAsLin2 identified 33 microbial pathways nominally associated with the intervention (exploratory FDR ≤ 0.25); one pathway (PWY-5104) also met the conventional FDR ≤ 0.10 threshold. Eighteen pathways were upregulated and 15 downregulated, with log fold changes ranging from −2.47 to 3.18. The 33 pathways were grouped into six functional clusters. L-alanine biosynthesis (PWY0-1061) was positively correlated with circulating IL-6 (ρ = 0.74; q < 0.01). Phosphopantothenate biosynthesis I (PANTO-PWY) was not significantly correlated with IL-6 (ρ = −0.46; q = 0.22).

**Conclusion:** Gut microbial functional analysis identified a positive association between L-alanine biosynthesis and circulating IL-6. This finding complements previously reported taxonomic shifts in this cohort and suggests a potential relationship between LCS consumption, microbial function, and host inflammatory signaling.

## 1. Introduction

Low-calorie sweeteners (LCS), including sucralose, acesulfame-potassium (Ace-K), aspartame, and saccharin, are consumed by about 41% of U.S. adults on a given day, primarily through intake of “diet” beverages (1). LCS provide sweetness with no or few calories; yet their role in cardiometabolic health remains controversial. Randomized controlled trials and prospective cohort studies often yield discordant findings, and a 2023 World Health Organization conditional recommendation advised against using LCS for weight management and prevention of noncommunicable diseases (2,3). A growing body of preclinical and clinical evidence suggests that LCS may be associated with compositional shifts in the gut microbiota (4,5), the community of microorganisms colonizing the gastrointestinal tract that are increasingly recognized as key modulators of cardiometabolic health. In a landmark human trial, Suez et al.(4) demonstrated that saccharin and sucralose impaired glycemic responses through individualized, microbiome-mediated mechanisms, with causality confirmed by fecal transplantation into germ-free mice. Large prospective cohort data further link intake of Ace-K, aspartame, and sucralose, the same compounds examined in the present study, to elevated type 2 diabetes risk (5).

Recent randomized trials demonstrate that replacing dietary sugars with sucralose reduced gut microbial alpha diversity over a 12-week exposure period (6). These compositional observations are of potential clinical relevance because Enterobacteriaceae have been linked to a pro-inflammatory gut environment, whereas Lactobacillales are associated with colonization resistance and immune regulation. Depletion of those protective functions may compromise gut barrier defenses (7). However, prior work did not characterize functional microbial metabolic pathway activity, nor did it examine associations between microbiome changes and circulating cardiometabolic biomarkers. Gut microbial functional profiling, enabled by shotgun whole-genome metagenomics and tools such as HUMAnN2 (8), provides direct quantification of microbial metabolic pathway capacity. That measure is orthogonal to taxonomic composition, and it is often more clinically informative than composition alone (8,9). Pathways involved in amino acid biosynthesis (10), peptidoglycan synthesis (11), short-chain fatty acid (SCFA) production (12), and vitamin metabolism (13) are of particular interest in cardiometabolic disease research. They offer potential mechanistic context for linking microbiome shifts to systemic inflammation and metabolic function (10)

Overweight women are of particular interest because they commonly exhibit low-grade systemic nflammation (14). That state is characterized by elevated circulating IL-6, leptin, and hs-CRP, markers that may be influenced by LCS-related changes in gut microbial function(7). The objectives of this secondary analysis were to: (1) to characterize gut microbial functional pathway profiles by visit (pre-vs. post-intervention) using shotgun DNA metagenomics and HUMAnN2 pathway profiling; and (2) to evaluate associations between visit-associated (i.e., pre-vs. post-intervention) microbial functional pathways and cardiometabolic biomarkers (IL-6, leptin, hs-CRP, HOMA-IR) among young overweight adult women who were enrolled in a prior pilot study investigating metabolic and health effects of 8 weeks of LCS intake (15).

## 2. Methods

### 2.1 Study Design and Participants

This is a secondary analysis of data from a single-arm, pre- and post-intervention pilot study, previously described elsewhere (15), including participant baseline characteristics, recruitment, and original analyses. Briefly, eight overweight (BMI ≥ 25 kg/m^2^) young (ages 18–35 years) adult women were enrolled in the longitudinal study. Participants were non-habitual LCS consumers and were instructed to consume 12 oz of caffeine-free Diet Pepsi, sweetened with sucralose and Ace-K, three times daily for eight weeks. We collected stool and fasting blood samples at baseline (Visit 1, pre-intervention) and at week 8 (Visit 2, post-intervention). The George Washington University Institutional Review Board (IRB #111635) approved the study, and it was registered on ClinicalTrials.gov (NCT03125356). All participants provided written informed consent.

### 2.2 Functional Pathway Profiling

We obtained microbiome sequencing data from the original study deposited in NCBI BioProject PRJNA995034. Briefly, stool samples were collected from eight participants, processed for DNA extraction (15), and extracted DNA was then sequenced using shotgun metagenomics on an Illumina NextSeq 500 platform. Functional pathway profiling was performed in the sequenced data using HUMAnN2 (v2.8) (8) with the UniRef90 protein database. We extracted pathway relative abundance data for downstream analysis, both unstratified (community-level) and stratified by species. This analysis used a linear mixed-model framework and the Benjamini-Hochberg procedure to control the false discovery rate (FDR; threshold ≤ 0.25). We then used a two-stage pathway clustering and biomarker-correlation approach to systematically examine pathway and cardiometabolic biomarker associations.

### 2.3 Cardiometabolic Biomarkers

Cardiometabolic biomarker analysis was performed using standard clinical chemistry and immunoassay methods by LabCorp (Burlington, NC) on an automated platform. Biomarkers measured at both visits included high-sensitivity C-reactive protein (hs-CRP), interleukin-6 (IL-6), leptin, adiponectin, TNF-α, fasting glucose, insulin, HOMA-IR, and triglycerides.

Biomarker data were normalized and clustered using HeliClust2.0 v2.0 (16). PCA was performed to visualize variation in metabolic profiles across time points (17).

### 2.4 Statistical Analysis

We tested associations among microbial pathways, cardiometabolic biomarkers, and host metadata (visit, age, race, BMI) with MaAsLin2 v1.26 (18) using a linear mixed model framework. Before testing, we filtered pathways to those present in at least 10% of samples. We fit each model on log_2_-transformed relative abundance, with standardized (z-scored) continuous fixed effects and a participant random intercept to account for the paired, repeated-measures design. We applied no additional normalization, since HUMAnN2 output is already expressed as relative abundance. Within each model, p values were adjusted using the Benjamini-Hochberg (BH) method.

Two fixed-effect specifications were fitted.

- Model A took the form: pathway ∼ Visit + Insulin + HOMA-IR + IL-6 + Leptin + Adiponectin + TNF-alpha + hs-CRP + Triglycerides + HDL + Sucrose + (1 | participant).
- Model B took the form: pathway ∼ Visit + Age + Race + BMI + (1 | participant).

Model A served as the primary visit-to-pathway screen. It supplied the pathways carried forward into the clustering and correlation analyses at a BH threshold of 0.25. Model B supplied the demographic and BMI results reported in Section 3.1 and the *Alistipes putredinis* pathway–BMI results reported in Section 3.2, at a BH threshold of 0.10. Model A included fasting glucose and insulin-adjusted dietary fiber, but the software dropped them due to collinearity.

We computed Spearman correlations between pathway cluster representatives and cardiometabolic biomarkers across all 16 samples. Given the pilot nature of this study (n = 16), we also report associations with nominal p < 0.05 as hypothesis-generating. At the full, unclustered scale (33 pathways × 12 biomarkers), a 1,000-permutation empirical FDR for the association between PWY0-1061 and IL-6 was q = 0.24. After reduction to six cluster representatives and three pre-specified biomarkers, the same association passed per-biomarker BH correction (q < 0.01). The contrast illustrates the multiple-testing burden that motivated the clustering step. We performed all analyses in R (v4.3.0).

## 3. Results

### 3.1 Participant Characteristics and Post-Intervention Cardiometabolic Profile

Eight young adult women (mean age 21.1 ± 1.7 years; mean body mass index 29.9 ± 2.8 kg/m^2^; 50% non-Hispanic White, 25% non-Hispanic Black, 12.5% Asian, 12.5% Other) completed the 8-week Diet Pepsi intervention (Table 1). We analyzed the post-intervention cardiometabolic biomarker pattern of participants. The PCA revealed that PC1 explained variance increased from 37.4% to 47.5%, while PC2 explained variance decreased from 26.9% to 21.7% (Figure 1). This indicates a shift toward a more dominant post-intervention metabolic pattern. Hierarchical clustering of the biomarker distance matrix showed changes in participant cluster membership between visits. Cardiometabolic biomarker analysis identified hs-CRP as significantly higher in participants of Other racial and ethnic backgrounds relative to White participants (coef = 2.12, SE = 0.23, q < 0.001). Leptin was significantly and positively associated with BMI (coef = 0.98, SE = 0.18, q = 0.0073), and hs-CRP showed a significant positive association with BMI (coef = 0.37, SE = 0.09, q = 0.0433).

**Table 1.** Participant characteristics at enrollment (n = 8).

| **Characteristic** | **(n = 8)** |
|---|---|
| **Sex** | 8 female (100%) |
| **Age, years** | 21.1 ± 1.7 |
| **BMI, kg/m<sup>2</sup>** | 29.9 ± 2.8 |
| **Non-Hispanic White** | 4 (50%) |
| **Non-Hispanic Black** | 2 (25%) |
| **Asian** | 1 (12.5%) |
| **Other** | 1 (12.5%) |
| **Total energy intake, kcal** | 1449.3 ± 261.5 |
| **Dietary fiber, g** | 14.5 ± 6.5 |

Values are mean ± SD unless otherwise specified.

**Figure 1.**
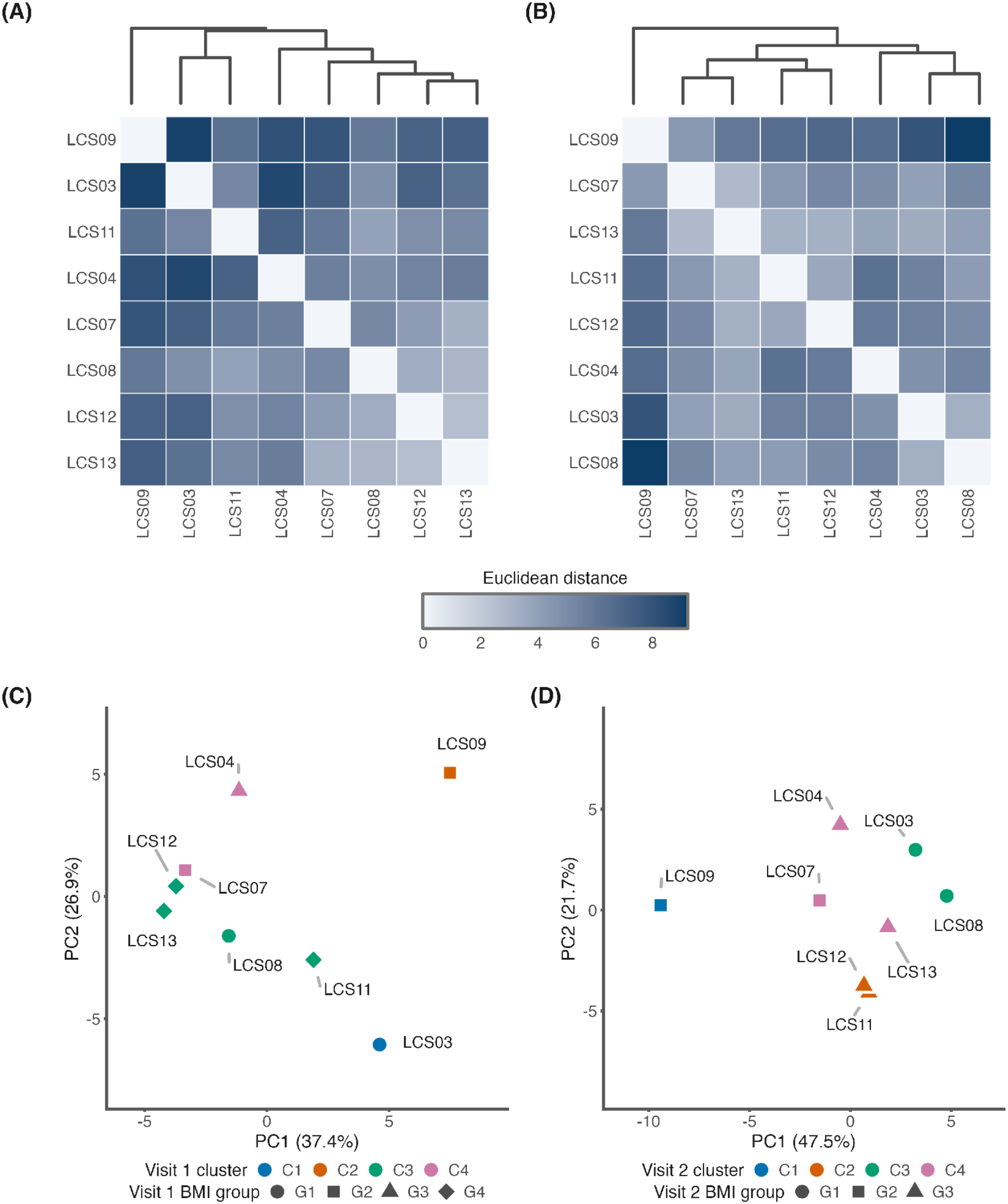
PCA and Distance Matrix Clustering Pre- and Post-Intervention. (A) Distance matrix heatmap with hierarchical clustering of metabolic biomarker profiles at pre-intervention (Visit 1, n = 8). (B) Distance matrix heatmap at post-intervention (Visit 2, n = 8), showing shifts in participant cluster membership. (C–D) PCA plots showing sample distribution at pre-intervention (PC1 37.4%, PC2 26.9%) and post-intervention (PC1 47.5%, PC2 21.7%). Point shapes represent BMI groups; colors represent clusters.

### 3.2 Microbial Functional Pathway Analysis

MaAsLin2 analysis identified 33 microbial functional pathways nominally associated with timepoint (pre vs. post-intervention) at an exploratory FDR ≤ 0.25 threshold. One pathway, PWY-5104, L-isoleucine biosynthesis IV, additionally met the conventional FDR ≤ 0.10 threshold (coef = 2.26, q = 0.095). Of the 33 pathways, 18 were upregulated, and 15 were downregulated post-intervention, with log fold changes ranging from −2.47 to 3.18 (Figure 2). Pathway direction was strongly taxon-dependent. All seven pathways attributed to *Akkermansia muciniphila* and all four arginine and ornithine biosynthesis pathways were downregulated. Coenzyme A biosynthesis pathways attributed to *Streptococcus parasanguinis* were upregulated. Pyrimidine deoxyribonucleotide pathways were mixed in direction depending on the contributing taxon: downregulated when *Akkermansia*-attributed, upregulated when *Haemophilus parainfluenzae*-attributed or community-level.

**Figure 2.**
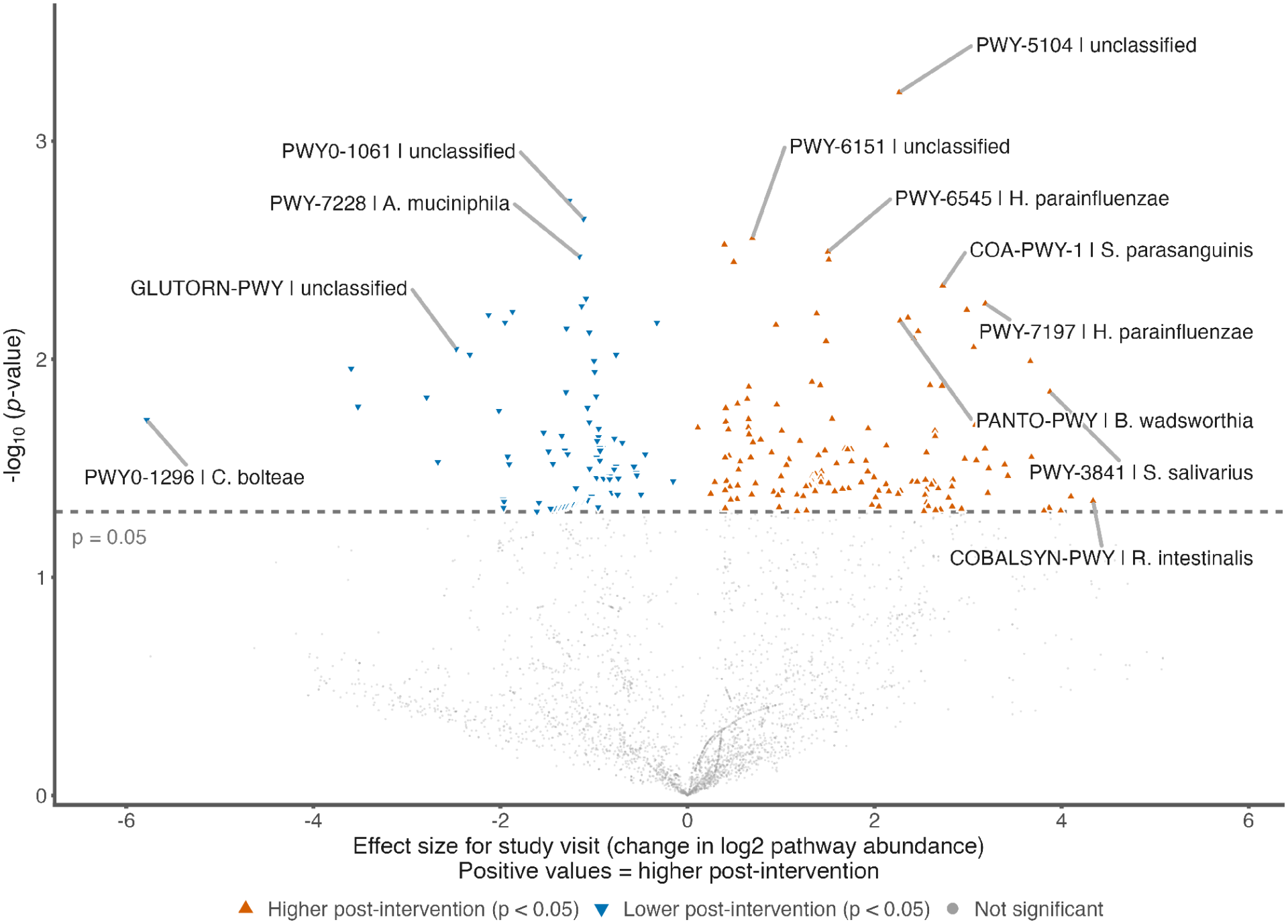
Volcano plot of visit-nominally associated microbial functional pathways. Volcano plot of all tested microbial metabolic pathways for their association with visit (pre vs. post intervention). The x-axis shows the MaAsLin2 LMM effect size (coefficient); the y-axis shows −log10(p-value). Blue points indicate pathways nominally upregulated post-intervention (p < 0.05); red points indicate nominally downregulated pathways.

We also observed race and BMI associations for pathways attributed to specific taxa. Functional pathways attributed to *Alistipes putredinis* were negatively associated with BMI. These included phosphopantothenate biosynthesis (PANTO-PWY; coef = −1.06, q < 0.001), nucleotide biosynthesis pathways (UMP biosynthesis, guanosine and adenosine ribonucleotide synthesis; coef range −0.97 to −1.06), and peptidoglycan biosynthesis pathways. The pattern suggests that higher adiposity is accompanied by a lower functional contribution from *A. putredinis*.

Multiple pathways were nominally enriched in specific racial and ethnic groups, particularly in the single Asian participant and the single participant classified as Other relative to White participants (n = 4). Because these two groups comprised one individual each, the pattern is confounded with participant identity and is reported descriptively rather than as evidence of a race- or ethnicity-associated effect.

### 3.3 Pathway–Cardiometabolic Biomarker Correlations

We retained pathways nominally associated with Visit at an exploratory FDR ≤ 0.25 threshold (BH-adjusted) for pathway and biomarker association analysis; we identify the subset that also meets the conventional FDR ≤ 0.10 threshold in Results. Correlating 33 such pathways against 12 biomarkers would have meant 396 tests. To manage this multiple-testing burden, pathways were grouped into six functional clusters by hierarchical clustering of their Spearman ρ correlation profiles across all 12 biomarkers (Ward.D2 linkage). We selected the pathway with the highest maximum |ρ| within each cluster as the cluster representative. This was reduced to 18 hypothesis tests (6 representatives × 3 pre-specified biomarkers: IL-6, Leptin, HOMA-IR) (Figure 3). We applied a per-biomarker Benjamini-Hochberg FDR correction to the six p-values per biomarker. The community-level superpathway of L-alanine biosynthesis (PWY0-1061 | unclassified) showed a significant positive correlation with circulating IL-6 (ρ = 0.74; q < 0.01). It was the only pathway-biomarker association to survive per-biomarker FDR correction.

**Figure 3.**
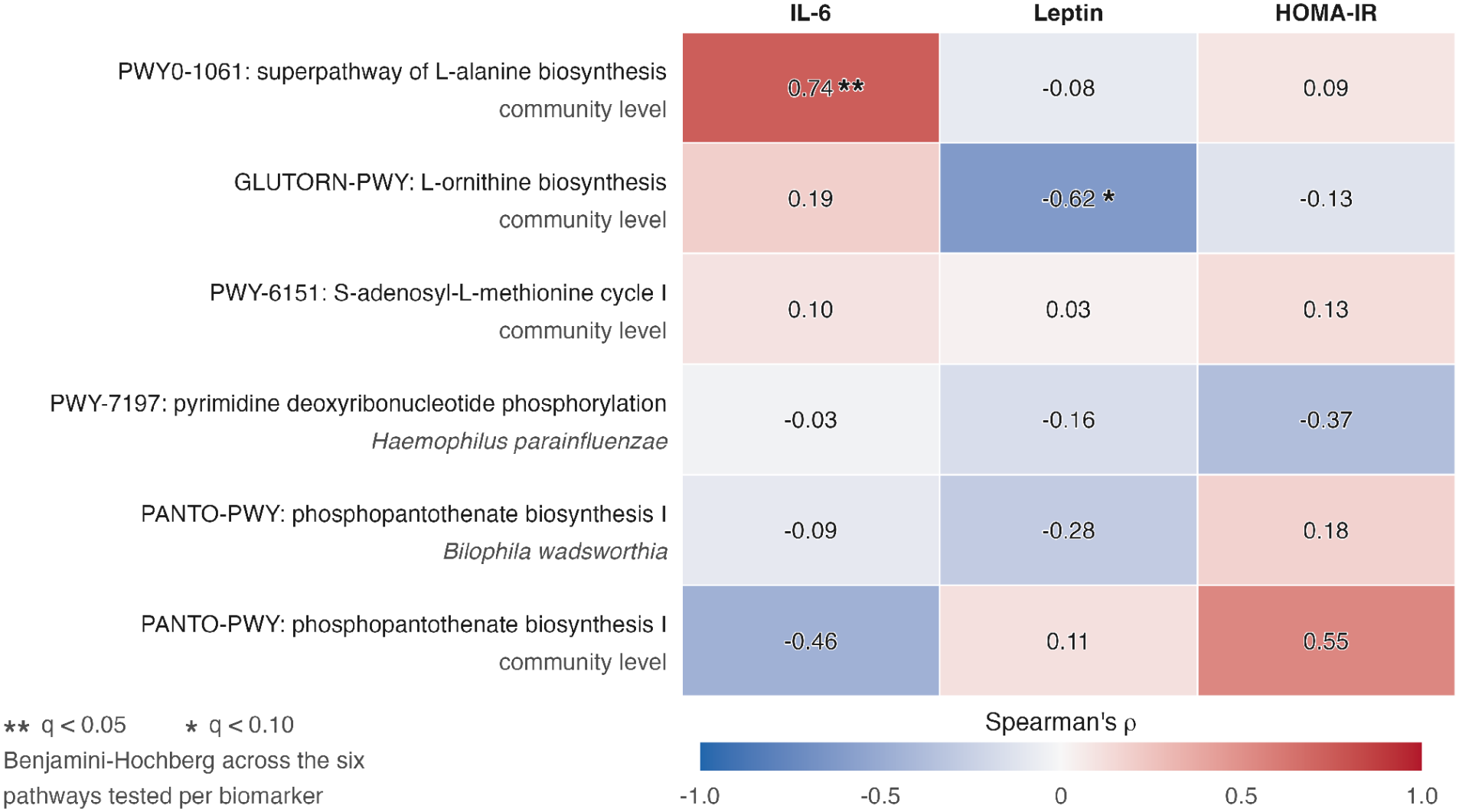
Spearman Correlation Heatmap: Pathway Cluster Representatives vs. Cardiometabolic Biomarkers. Spearman ρ correlation heatmap between six visit-significant pathway cluster representatives and three pre-specified cardiometabolic biomarkers (IL-6, Leptin, HOMA-IR) across all 16 samples (8 participants × 2 visits). Cell values show Spearman ρ. ** q < 0.05 (per-biomarker BH FDR); * q < 0.10. Red = positive association; blue = negative association.

Phosphopantothenate biosynthesis I (PANTO-PWY | unclassified) was not significantly correlated with IL-6 (ρ = −0.46; q = 0.22) but showed a positive association with HOMA-IR (ρ = 0.55, p = 0.028; q = 0.17). L-ornithine biosynthesis (GLUTORN-PWY | unclassified) showed an inverse association with leptin (ρ = −0.62, p = 0.011; q = 0.064). These secondary associations were attenuated following FDR correction and are reported as hypothesis-generating.

## 4. Discussion

This secondary analysis identified 33 gut microbial functional pathways nominally associated with 8 weeks of LCS-sweetened diet soda consumption at an exploratory FDR ≤ 0.25 threshold. One of these, PWY-5104 (L-isoleucine biosynthesis IV), also met the conventional FDR ≤ 0.10 threshold. Among the 33 identified functional pathways, the community-level superpathway of L-alanine biosynthesis (PWY0-1061) was significantly and positively correlated with circulating IL-6 (ρ = 0.74; q < 0.01). It was the only pathway-biomarker association to survive FDR correction. This functional pathway-level analysis confirms the taxonomic observations of the parent study (15) toward a potential link between gut microbial functional activity and systemic inflammation markers. Given the small sample size and single-arm design, caution is needed in interpreting the findings, which are preliminary and hypothesis-generating.

The association between PWY0-1061 and IL-6 has a biologically plausible, though unestablished, mechanistic basis. L-alanine is the first amino acid incorporated into the bacterial peptidoglycan stem peptide, so higher microbial L-alanine biosynthetic capacity is consistent with greater peptidoglycan production and cell wall turnover (19). Fragments of peptidoglycan, known as muropeptides, can cross the intestinal epithelium and bind the pattern recognition receptors NOD1 and NOD2 on host immune cells. This binding activates NF-κB and MAPK signaling, which drives macrophage production of IL-6 and TNF-α (9,20). Greater microbial capacity for L-alanine biosynthesis could therefore plausibly increase muropeptide shedding and NOD1/NOD2 engagement, offering one route by which the PWY0-1061 signal could relate to higher circulating IL-6. In inflammatory bowel disease cohorts. The PWY0-1061 pathway has been found to be enriched in Enterobacteriaceae-dominated gut communities (21).

The PWY0-1061 signal in this dataset was detected at the community (unclassified) level, indicating that multiple taxa, including confirmed contributors *Citrobacter freundii, Escherichia coli, Haemophilus parainfluenzae, Streptococcus australis*, and *Streptococcus parasanguinis*, contributed collectively to this functional output, rather than a single dominant species. This is consistent with a pathway-level rather than organism-specific association: microbial function is often more conserved across communities than the specific taxa that carry it out, since unrelated species can encode the same metabolic pathway, and a function distributed across several taxa can be more biologically meaningful than one traced to a single organism (22). Whether this pattern reflects a response to the LCS intervention, pre-existing individual differences in microbiome function, or both cannot be determined from these data alone.

The association between PWY0-1061 and IL-6 observed here is consistent with the taxonomic pattern previously reported (15): increased relative abundance of Gammaproteobacteria, specifically Enterobacteriaceae, alongside decreased Lactobacillales, following 8 weeks of sucralose- and Ace-K-containing diet soda consumption. Enterobacteriaceae are known to constitutively carry the enzymatic genes underlying PWY0-1061, including alanine racemase and alanine aminotransferases. Lactobacillales, in contrast, are associated with competitive suppression of Enterobacteriaceae through lactic acid production (7). This study observed both a nominal shift toward Enterobacteriaceae and elevated community-level L-alanine biosynthetic capacity. That co-occurrence is consistent with prior literature reporting that LCS exposure, particularly sucralose, is associated with Proteobacteria enrichment in both animal and human studies (15,23,24).

The inverse, though non-significant, association between PANTO-PWY and IL-6 (ρ = −0.46) adds contextual interest. Phosphopantothenate biosynthesis, i.e., vitamin B5 synthesis, is a precursor step in coenzyme A (CoA) biosynthesis. CoA is required for SCFA production and lipid metabolism, processes associated with anti-inflammatory gut homeostasis (25). The functional capacity for phosphopantothenate biosynthesis was negatively associated with BMI in the *A. putredinis*-attributed pathways in this cohort (coef ≈ −1.06, q < 0.001). Two additional secondary, non-significant associations support this inverse pattern. PANTO-PWY also showed a nominal positive association with HOMA-IR (ρ = 0.55, p = 0.028; q = 0.17). L-ornithine biosynthesis (GLUTORN-PWY), also detected at the community (unclassified) level, showed a nominal inverse association with leptin (ρ = −0.62, p = 0.011; q = 0.064). Taken together, these nominal associations suggest that the same shift toward Enterobacteriaceae-dominated, pro-inflammatory community function, reflected in elevated PWY0-1061, may coincide with reduced *Alistipes*-associated, anti-inflammatory metabolic capacity. None of these secondary associations survived FDR correction, so they should be interpreted cautiously as hypothesis-generating only.

The main limitation of this study was the relatively small sample size (N=8). This limited the statistical power to detect modest associations. Also, because shotgun DNA metagenomic sequencing directly assesses microbial gene content and functional potential, but it does not determine whether these genes are actively expressed or contribute to metabolic activity. The observed functional associations therefore require validation through complementary approaches such as transcriptomics, proteomics, and/or metabolomics. The study also has important strengths. The pathway-clustering approach provided a principled strategy for reducing dimensionality and managing multiple-testing burden in a small-sample pilot setting. In addition, prior evidence linking peptidoglycan-associated pathways to host immune and inflammatory signaling supports the biological plausibility of the primary finding: the association between PWY0-1061 and IL-6.

## 5. Conclusion

Functional analysis of the gut microbiome revealed that L-alanine biosynthesis is positively correlated with circulating IL-6. This was observed in a cohort of young overweight adult women who consumed LCS-sweetened diet soda for 8 weeks. This finding is consistent with the prior taxonomic shifts reported in this cohort and suggests a potential link between inflammatory signaling and LCS consumption. Larger trials with longer LCS exposure durations (e.g., 6 months) will provide greater clarity on the effects of LCS on microbial functional features and their relationship with cardiometabolic biomarkers.

## Data Availability

All data produced in the present study are available upon reasonable request to the authors

## Data Availability Statement

Sequence data from the parent study are deposited in the NCBI Sequence Read Archive (accession number SUB13028274). Additional data analyzed in this secondary analysis are available upon reasonable request from the corresponding authors.

## Ethics Statement

This secondary analysis used data from a study conducted in accordance with the Declaration of Helsinki. The parent study received ethical approval from the George Washington University Institutional Review Board (IRB #111635). All participants provided written informed consent prior to enrollment in the original study.

## Author Contributions

PA: Formal Analysis, Methodology, Writing—original draft.

AC: Conceptualization, Investigation, Data Curation, Formal Analysis, Methodology, and Funding Acquisition.

CO: Project Administration, Supervision, Writing—original draft, Writing—review and editing. MP-L: Writing—review and editing.

KC: Investigation, Data Curation, Formal Analysis, and Methodology.

## Acknowledgments

The authors acknowledge the participants of the original study (Sylvetsky et al., 2024). Secondary analyses were conducted using resources provided by the George Washington University computational infrastructure.

## References

1. Sylvetsky AC, Jin Y, Clark EJ, Welsh JA, Rother KI, Talegawkar SA. Consumption of Low-Calorie Sweeteners among Children and Adults in the United States. Journal of the Academy of Nutrition and Dietetics. 2017 Mar 1;117(3):441–448.e2. doi:10.1016/j.jand.2016.11.004

2. Sylvetsky AC, Rother KI. Nonnutritive Sweeteners in Weight Management and Chronic Disease: A Review. Obesity (Silver Spring). 2018 Apr;26(4):635–40. doi:10.1002/oby.22139 PubMed PMID: 29570245; PubMed Central PMCID: PMC5868411.

3. Khan TA, Ayoub-Charette S, Sievenpiper JL. Non-nutritive Sweeteners and Health: Reconciling Evidence and Interrogating Guideline Disconnects. Adv Nutr. 2024 Dec;15(12):100328. doi:10.1016/j.advnut.2024.100328 PubMed PMID: 39675839; PubMed Central PMCID: PMC11705572.

4. Suez J, Cohen Y, Valdés-Mas R, Mor U, Dori-Bachash M, Federici S, et al. Personalized microbiome-driven effects of non-nutritive sweeteners on human glucose tolerance. Cell. 2022 Sep 1;185(18):3307–3328.e19. doi:10.1016/j.cell.2022.07.016 PubMed PMID: 35987213.

5. Debras C, Deschasaux-Tanguy M, Chazelas E, Sellem L, Druesne-Pecollo N, Esseddik Y, et al. Artificial Sweeteners and Risk of Type 2 Diabetes in the Prospective NutriNet-Santé Cohort. Diabetes Care. 2023 Sep 1;46(9):1681–90. doi:10.2337/dc23-0206 PubMed PMID: 37490630; PubMed Central PMCID: PMC10465821.

6. Haslam DE, Abirami K, Starr JR, Unnikrishnan R, Lasky-Su J, Gayathri R, et al. Effect of Replacing Added Sugars with Sucralose on Gut Microbiome Composition Among Asian Indian Adults in Two 12-week Randomized Controlled Trials. Curr Dev Nutr. 2025 Nov 12;9(12):107600. doi:10.1016/j.cdnut.2025.107600 PubMed PMID: 41451263; PubMed Central PMCID: PMC12731820.

7. Conz A, Salmona M, Diomede L. Effect of Non-Nutritive Sweeteners on the Gut Microbiota. Nutrients. 2023 Apr 13;15(8):1869. doi:10.3390/nu15081869 PubMed PMID: 37111090; PubMed Central PMCID: PMC10144565.

8. Franzosa EA, McIver LJ, Rahnavard G, Thompson LR, Schirmer M, Weingart G, et al. Species-level functional profiling of metagenomes and metatranscriptomes. Nat Methods. 2018 Nov;15(11):962–8. doi:10.1038/s41592-018-0176-y PubMed PMID: 30377376; PubMed Central PMCID: PMC6235447.

9. Clarke TB, Davis KM, Lysenko ES, Zhou AY, Yu Y, Weiser JN. Recognition of peptidoglycan from the microbiota by Nod1 enhances systemic innate immunity. Nat Med. 2010 Feb;16(2):228–31. doi:10.1038/nm.2087 PubMed PMID: 20081863; PubMed Central PMCID: PMC4497535.

10. Boulangé CL, Neves AL, Chilloux J, Nicholson JK, Dumas ME. Impact of the gut microbiota on inflammation, obesity, and metabolic disease. Genome Med. 2016 Apr 20;8(1):42. doi:10.1186/s13073-016-0303-2 PubMed PMID: 27098727; PubMed Central PMCID: PMC4839080.

11. Pedersen HK, Gudmundsdottir V, Nielsen HB, Hyotylainen T, Nielsen T, Jensen BAH, et al. Human gut microbes impact host serum metabolome and insulin sensitivity. Nature. 2016 Jul 21;535(7612):376–81. doi:10.1038/nature18646 PubMed PMID: 27409811.

12. Rodrigues E-Lacerda R, Fang H, Robin N, Bhatwa A, Marko DM, Schertzer JD. Microbiota and Nod-like receptors balance inflammation and metabolism during obesity and diabetes. Biomed J. 2023 Oct;46(5):100610. doi:10.1016/j.bj.2023.100610 PubMed PMID: 37263539; PubMed Central PMCID: PMC10505681.

13. Böttcher J, Sibon OCM, El Aidy S. Coenzyme A metabolism: a key driver of gut microbiota dynamics and metabolic profiles. FEMS Microbiol Rev. 2025 Oct 11;49:fuaf051. doi:10.1093/femsre/fuaf051 PubMed PMID: 41074819; PubMed Central PMCID: PMC12570885.

14. Khanna D, Khanna S, Khanna P, Kahar P, Patel BM. Obesity: A Chronic Low-Grade Inflammation and Its Markers. Cureus. 2022 Feb;14(2):e22711. doi:10.7759/cureus.22711 PubMed PMID: 35386146; PubMed Central PMCID: PMC8967417.

15. Sylvetsky AC, Clement RA, Stearrett N, Issa NT, Dore FJ, Mazumder R, et al. Consumption of sucralose- and acesulfame-potassium-containing diet soda alters the relative abundance of microbial taxa at the species level: findings of two pilot studies. Appl Physiol Nutr Metab. 2024 Jan 1;49(1):125–34. doi:10.1139/apnm-2022-0471 PubMed PMID: 37902107.

16. Appiah PY. HeliClust2.0: Hierarchical Clustering for Metabolomic and Microbiome Sequence Data [Internet]. 2026 Jul 10.doi:10.5281/zenodo.21287626

17. Rahnavard A, Chatterjee S, Sayoldin B, Crandall KA, Tekola-Ayele F, Mallick H. Omics community detection using multi-resolution clustering. Bioinformatics. 2021 Oct 25;37(20):3588–94. doi:10.1093/bioinformatics/btab317 PubMed PMID: 33974004; PubMed Central PMCID: PMC8545346.

18. Mallick H, Rahnavard A, McIver LJ, Ma S, Zhang Y, Nguyen LH, et al. Multivariable association discovery in population-scale meta-omics studies. PLoS Comput Biol. 2021 Nov;17(11):e1009442. doi:10.1371/journal.pcbi.1009442 PubMed PMID: 34784344; PubMed Central PMCID: PMC8714082.

19. Rojas ER, Huang KC. Regulation of microbial growth by turgor pressure. Curr Opin Microbiol. 2018 Apr;42:62–70. doi:10.1016/j.mib.2017.10.015 PubMed PMID: 29125939.

20. Caruso R, Lo BC, Núñez G. Host-microbiota interactions in inflammatory bowel disease. Nat Rev Immunol. 2020 Jul;20(7):411–26. doi:10.1038/s41577-019-0268-7 PubMed PMID: 32005980.

21. Dubinsky V, Reshef L, Bar N, Keizer D, Golan N, Rabinowitz K, et al. Predominantly Antibiotic-resistant Intestinal Microbiome Persists in Patients With Pouchitis Who Respond to Antibiotic Therapy. Gastroenterology. 2020 Feb;158(3):610–624.e13. doi:10.1053/j.gastro.2019.10.001 PubMed PMID: 31605691.

22. Langille MGI. Exploring Linkages between Taxonomic and Functional Profiles of the Human Microbiome. mSystems. 2018 Mar 27;3(2):10.1128/msystems.00163-17. doi:10.1128/msystems.00163-17

23. Suez J, Korem T, Zeevi D, Zilberman-Schapira G, Thaiss CA, Maza O, et al. Artificial sweeteners induce glucose intolerance by altering the gut microbiota. Nature. 2014 Oct 9;514(7521):181–6. doi:10.1038/nature13793 PubMed PMID: 25231862.

24. Suez J, Korem T, Zilberman-Schapira G, Segal E, Elinav E. Non-caloric artificial sweeteners and the microbiome: findings and challenges. Gut Microbes. 2015;6(2):149–55. doi:10.1080/19490976.2015.1017700 PubMed PMID: 25831243; PubMed Central PMCID: PMC4615743.

25. Patra D, Banerjee D, Ramprasad P, Roy S, Pal D, Dasgupta S. Recent insights of obesity-induced gut and adipose tissue dysbiosis in type 2 diabetes. Front Mol Biosci. 2023 Sep 28;10. doi:10.3389/fmolb.2023.1224982

